# Blood Lead Levels and Alzheimer’s Disease Mortality in NHANES: Addressing Temporal Confounding Through a Study Exit Covariate

**DOI:** 10.1101/2024.07.20.24310751

**Authors:** Aaron Grossman

## Abstract

**Background:** The relationship between blood lead levels (BLLs) and Alzheimer’s disease (AD) mortality remains unclear. Prior efforts are likely hampered by structural temporal confounding. In pooled historical cohorts with fixed mortality-linked ascertainment windows, differences in enrollment timing dictate individual follow-up duration, disproportionately concentrating cumulative person-time and advanced age within earlier cohorts. Crucially, across these same historical periods, environmental lead exposure steeply declined while clinical recognition and cause-of-death certification of AD expanded. This systematic imbalance forces earlier, higher-exposed cohorts to contribute more person-time during the modern high-ascertainment era, while later cohorts are right-censored prematurely. This confounds exposure with diagnostic landscape, biasing conventional survival estimates toward the null.

**Methods:** In a two-phase design, we first replicated a prior Continuous NHANES (1999–2008) cohort with updated mortality follow-up through December 2019 (Phase 1), then expanded the sample by pooling Continuous NHANES with NHANES III (1988–1994) to evaluate 21,308 participants aged ≥40 years with 350 AD deaths (Phase 2). To correct for temporal confounding, we compared conventional Cox models with and without conditioning on a structural covariate: calendar time at study exit.

**Results:** Phase 1 replication with extended follow-up neutralized the previously reported positive hazard ratio to an attenuated null (HR=1.00; 95% CI: 0.78–1.28). In Phase 2 pooled analyses, conventional Cox models also showed no meaningful overall association (HR=0.86; 95% CI: 0.70–1.05). However, incorporating the study exit covariate yielded a robust inverse association (HR=0.57; 95% CI: 0.46–0.70), which remained stable across categorical specifications and sensitivity analyses excluding early deaths (10-year exclusion: HR=0.54; 95% CI: 0.42–0.69). Exit-year stratified analyses revealed consistent within-stratum inverse associations, consistent with Simpson’s paradox-type aggregation effects.

**Conclusions:** Systematic differences in enrollment-driven person-time across evolving diagnostic eras profoundly distort exposure-disease relationships in long-latency historical cohorts. Conditioning on calendar time at study exit provides a method for equalizing the ascertainment landscape across heterogeneous study periods without restricting the sample or compromising model structure. However, the observed inverse BLL–AD association should not be interpreted as evidence of a protective effect of lead exposure. Future studies are needed to establish the true nature of this relationship.

## Introduction

Alzheimer’s disease (AD) is a progressive neurodegenerative disorder with an age-adjusted prevalence that has increased over recent decades.[1–3] Altered concentrations of several metals have been observed in AD brain tissue, suggesting that environmental exposures may influence disease pathogenesis.[4, 5] One such metal, lead (Pb), is a well-established neurotoxin with historically widespread exposure resulting from its use in gasoline, paint, plumbing, and industrial applications. Although lead exposure has been studied extensively in relation to cognitive decline, relatively few epidemiologic investigations have examined its relationship with AD.[6–8] A prior analysis of NHANES data by Horton et al. reported a positive, though nonsignificant, association between baseline blood lead level (BLL) and AD mortality.[6] A similar study by Wang et al. using Medicare-linked NHANES data found no association between blood lead and incident AD, though cumulative bone lead was associated with increased risk.[9]

Evaluating the Pb-AD relationship presents several methodological challenges. Lead exposure in the U.S. has declined dramatically following regulatory actions beginning in the 1970s.[10] In pooled analyses spanning multiple recruitment waves with a fixed mortality linkage window, this secular decline creates a structural imbalance: earlier-enrolled participants have both higher baseline blood lead level (BLL) and longer potential follow-up duration. Crucially, because AD mortality is only ascertainable at death, later-enrolled cohorts—who are more frequently right-censored as survivors—contribute person-time without opportunity for outcome detection, regardless of their true disease burden. This asymmetry is compounded by the increasing appearance of AD as an underlying cause of death on death certificates over the study period.[11] This means that earlier-enrolled, higher-exposed participants not only accumulate more follow-up time, but do so increasingly within calendar periods characterized by greater ascertainment of AD mortality. Together, these mechanisms may confound exposure with diagnostic era, biasing conventional survival estimates in ways that standard covariate adjustment, including adjustment for survey cycle or baseline calendar year, cannot resolve, since the confounding accrues continuously throughout follow-up rather than at enrollment.

Studies of long-latency diseases are particularly vulnerable to bias when environmental exposures change substantially over time. When exposure levels decline across recruitment cohorts while follow-up duration varies, conventional survival models may distort exposure–response relationships. To explore these issues, we reassessed the association between lead exposure and AD mortality using NHANES data in a two-phase design. Our objectives were to: (1) re-evaluate the previously reported BLL–AD mortality association within the 1999–2008 NHANES cohort using extended mortality follow-up;[6] (2) evaluate whether associations vary across survey cycles and calendar time; (3) assess whether accounting for a novel structural covariate alters the observed relationship; and (4) examine the impact of excluding early deaths to reduce potential reverse causality. We hypothesized that secular declines in lead exposure, combined with differential opportunities for outcome ascertainment across survey cycles, may obscure underlying exposure–response patterns.

## Methods

### Study overview

We conducted a two-phase study examining BLL and AD mortality. Phase 1 replicated Horton et al.’s Continuous NHANES (1999-2008) analysis using updated mortality data through 2019, then extended it by adjusting for temporal confounding. Phase 2 integrated Continuous NHANES with NHANES III (1988-1994) to increase statistical power and extend observation period.

### Data sources

NHANES comprises cross-sectional studies of the U.S. civilian noninstitutionalized population conducted by NCHS, with data released in two-year cycles from 1999 onward and periodically before that. Data include standardized questionnaires, physical examinations, and laboratory testing.[12, 13] The NCHS Ethics Review Board approved all protocols; adults provided written informed consent. Data were accessed January 2024.

### Exposure assessment

Blood samples were collected in NHANES mobile examination center visits. Whole blood specimens were processed, stored frozen at –30°C, and shipped to the Division of Laboratory Sciences at the National Center for Environmental Health, CDC, for analysis. Blood lead concentrations were measured by graphite furnace atomic absorption spectrophotometry in NHANES III (detection limit: 1.0 µg/dL) and inductively coupled plasma mass spectrometry in Continuous NHANES (detection limit: 0.3 µg/dL). Detailed laboratory methods and quality control procedures have been described elsewhere.[14, 15]

### Outcomes assessment

AD mortality was identified as the underlying cause of death using International Classification of Diseases codes (ICD-9) code 331.0 (pre-1999) and ICD-10 code G30 (1999 onward).[16, 17] We used updated NHANES-NDI linked mortality files through December 2019, which employ an improved probabilistic matching algorithm compared to the 2014 files used by Horton et al.[16, 17] Unmatched participants were assumed alive at end of follow-up.

### Phase 1: Continuous NHANES (1999-2008)

#### Replication analysis

We pooled Continuous NHANES data from five two-year cycles, 1999-2008 (hereafter, Cycles 1-5). The study population included participants aged ≥60 years with available BLL measurements. Covariates included age, sex, race/ethnicity, and poverty-to-income ratio (PIR <2, ≥2, or missing). Current smokers were identified by serum cotinine ≥10 ng/mL when available, or by self-report of cigarette, pipe, or cigar use.[18] BLL values below the detection limit (0.3 µg/dL) were initially imputed at 0.2 µg/dL to match Horton et al. We examined the continuous association between natural log-transformed BLL and AD mortality using Cox proportional hazards regression.

#### Modification analyses

After replication, we implemented modifications to address potential biases. We excluded participants with BLL below the detection limit as secular declines mean imputing with a single value would introduce systematic bias varying by cycle. We lowered the minimum age to 50 years at baseline to capture additional person-time and AD events given extended follow-up through 2019. We also excluded participants with top-coded ages at baseline (≥85 years in Cycles 1-4 and ≥80 years in Cycle 5), consistent with other NHANES analyses.[19] This ensures precise age adjustment in Cox models.

#### Covariates and Temporal Confounding Adjustment

Cox models included covariates for demographic and health factors associated with mortality risk: age, sex, race/ethnicity, education (<12 vs. ≥12 years), poverty-to-income ratio, current smoking status, and history of cardiovascular disease, cancer, hypertension, diabetes, and osteoporosis.

Pooling introduces temporal confounding arising from three interrelated features of these data. First, secular declines in environmental lead exposure cause participants recruited earlier (e.g., 1999–2000) to have higher baseline BLL and steeper subsequent declines than participants enrolled later (e.g., 2007–2008). Second, opportunity for AD mortality ascertainment differs across calendar time: earlier cohorts have longer possible follow-up, and within each cycle, participants who die later—or who remain alive at study end—have more limited opportunity for AD to be detected, because AD mortality is only ascertainable at death. Third, because earlier recruitment is associated with higher BLL, longer follow-up time becomes disproportionately concentrated among participants with higher exposure, potentially generating a spurious positive association between BLL and AD mortality that does not reflect a causal relationship. Adjusting for survey cycle or baseline calendar year does not resolve this, as such adjustments do not capture the continuous variation in both exposure trajectories and outcome ascertainment that accrues over follow-up.

To address this, we constructed a calendar-effect covariate representing approximate calendar time at study exit (Figure 1). For each participant, we calculated this by adding their follow-up duration to a cycle-specific offset, thereby aligning individuals by exit rather than entry year. These offsets were defined as: 0 months for Cycle 1 (1999–2000), 24 months for Cycle 2 (2001–2002), 48 months for Cycle 3 (2003–2004), 72 months for Cycle 4 (2005–2006), and 96 months for Cycle 5 (2007–2008), calculated as follows:

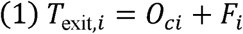

**Figure 1.**
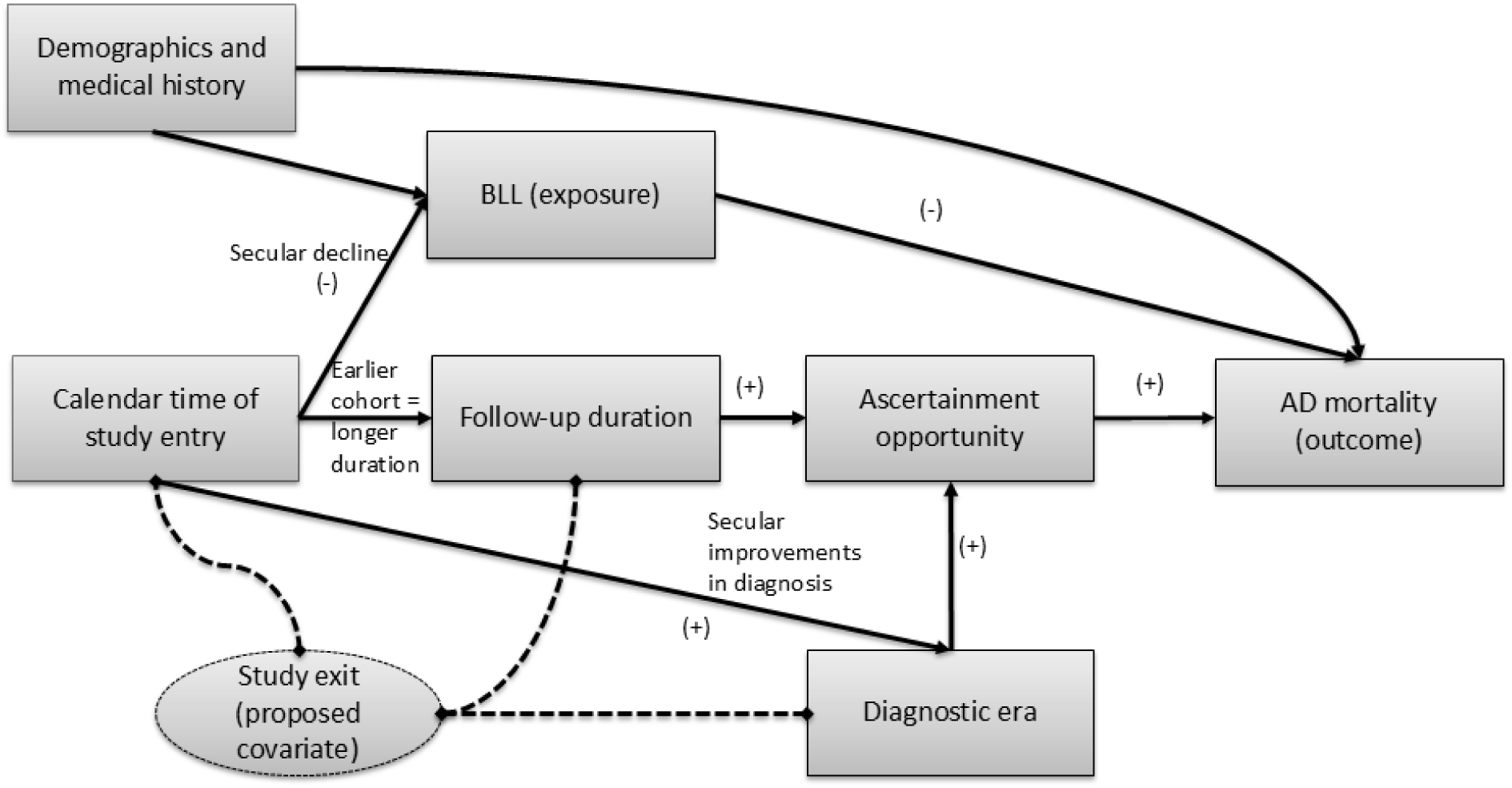
Directed Acyclic Graph (DAG) illustrating the structural and temporal confounding pathways between blood lead levels and Alzheimer’s disease mortality. The primary analytical pathway evaluates the hypothesis of an inverse association between baseline BLL and AD mortality. Standard measured covariates, including demographics and medical history, influence both baseline BLL and mortality risk. Temporal confounding is introduced by calendar time of study entry, which operates through three concurrent pathways: the secular decline in environmental lead exposure, which determines baseline BLL; follow-up duration, which is longer for earlier-enrolled cohorts; and secular improvements in AD diagnostic recognition and increasing use of AD as the underlying cause of death on death certificates. Together, the latter two pathways systematically increase the opportunity for AD mortality ascertainment among earlier-enrolled cohorts, independently of true disease burden. Because these pathways continue to operate throughout follow-up and cannot be resolved by adjusting for study entry calendar time alone, conditioning on study exit calendar time—a structural covariate that simultaneously indexes individual follow-up duration and the diagnostic era at the point of ascertainment—is proposed to substantially reduce temporal confounding arising from differential follow-up duration and diagnostic era, equalizing exposure trajectories and ascertainment opportunities across pooled cohorts.

Where *T*_exit,*i*_ is the study exit calendar time for participant *I; O_ci_* is the cycle-specific calendar offset for cohort *c* to which individual *i* belongs; and *F_i_* is individual follow-up duration in will have different *T*_exit_ values, and two participants with identical follow-up durations but months. Thus, two participants enrolled in the same cycle but with different follow-up durations enrolled in different cycles will also differ. This approach serves three purposes: it equalizes ascertainment opportunity across cohorts, accounts for secular declines in BLL experienced differentially across recruitment periods, and adjusts for period effects including changes in AD diagnosis and cause-of-death reporting. In pooled analyses, this calendar-effect covariate was included as a continuous predictor in Cox proportional hazard models.

### Phase 2: Combined NHANES datasets (1988-2008)

We next integrated Continuous NHANES (1999-2008) with NHANES III (1988-1994), including nonpregnant participants aged ≥40 years at baseline. This lower age threshold allows NHANES III participants to age into the at-risk period for AD during extended follow-up, contributing to both person-time and events. We excluded participants whose follow-up would extend beyond age 100 years, as NDI matching accuracy declines substantially at extreme ages. Because NHANES III used a higher detection limit (1.0 µg/dL), we harmonized datasets by excluding all participants with BLL <1.0 µg/dL. Participants with top-coded ages at baseline (≥85 years in Continuous NHANES Cycles 1-4, ≥80 years in Cycle 5, and ≥90 years in NHANES III) were excluded following cycle-specific NCHS confidentiality rules. Covariates included sex, race/ethnicity, education, poverty-to-income ratio (<1.30, 1.30-3.49, ≥3.50, and missing), current smoking status (using criteria from Phase 1), and medical history (cardiovascular disease, cancer excluding nonmelanoma skin cancer, hypertension, diabetes, and osteoporosis).

The calendar-effect covariate was recalibrated to span the full study period (1988–2019), with a reference point at March 1990. For each participant, we calculated time from March 1990 to the midpoint of their recruitment cycle, then added their individual follow-up time. Specifically, cycle midpoint offsets from the March 1990 reference were: 0 months for NHANES III Phase 1 (1988-1991), 24 months for NHANES III Phase 2 Years 4 (1992-1993), 42 months for NHANES III Phase 2 Years 5-6 (1993-1994), and 117, 141, 165, 189, and 213 months for Continuous NHANES Cycles 1-5 (1999-2000, 2001-2002, 2003-2004, 2005-2006, 2007-2008), respectively. This aligns all participants on a common calendar timeline from 1988 to 2019, accounting for differential exposure trajectories and outcome ascertainment across the full study period.

We evaluated dose-response using three approaches: continuous log-transformed BLL stratified by 5-year exit intervals; categorical BLL (1.0–2.4, 2.5–4.9, 5.0–7.4, 7.5–9.9, ≥10 µg/dL) with linear trend tests; and restricted cubic splines with five knots at the 5th, 27.5th, 50th, 72.5th, and 95th percentiles, with the reference at the 12.5th percentile. [20]

### Statistical analysis

Survival time was calculated from examination date to death or December 2019. Cox proportional hazards models estimated HRs and 95% CIs for AD mortality, treating non-AD deaths as censored. The fully adjusted model took the form:

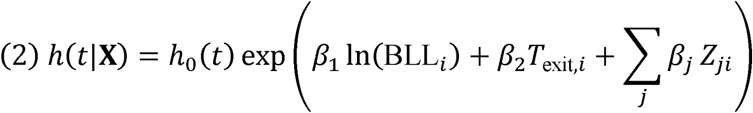

Where *h*(*t*|**X**) is the hazard of AD mortality at time *t* for individual *I*; *h*□(*t*) represents the baseline hazard function at time *t*; *β*□ represents the coefficient for natural log-transformed blood lead level (ln(BLL*i*)); *T_exit,i_*is as defined in Equation 1 and represents the specific calendar year participant *i* exited the cohort (either via all-cause mortality or administrative censoring); and *Z_ji_* represents the standard baseline covariates adjusted for in the model.

Although AD mortality represented a small proportion of the cohort, Cox model precision depends on the absolute number of events, not the percentage.[21] Our analysis included 350 AD deaths, providing adequate statistical power to detect associations. Proportional hazards assumptions were verified using Schoenfeld residuals. Analyses were conducted in SAS® OnDemand for Academics without survey weights, as our focus was internal validity rather than population-representative estimates. Sensitivity analyses excluded participants dying within 5 or 10 years of examination to minimize reverse causation.[22, 23] In addition, to evaluate the potential influence of competing mortality, we performed a post-hoc analysis comparing mean age at death from non-AD causes across BLL categories.

## Results

### Phase 1: NHANES 1999-2008

Participant characteristics by Continuous NHANES cycle and overall are presented in Table 1. The replication sample (n=8,080) matched the Horton et al. study population,[6] but extended mortality follow-up (through December 2019) and improved NDI matching algorithms substantially increased ascertained deaths (from 81 [1.00%] to 231 [2.86%]).

**Table 1.** Demographic and clinical characteristics of Continuous NHANES study participants by cycle (1999-2008).

| Variable | Overall | NHANES Cycle |  |  |  |  | P-value * |
| --- | --- | --- | --- | --- | --- | --- | --- |
|  |  | 1999-2000<br>(1) | 2001-2002<br>(2) | 2003-2004<br>(3) | 2005-2006<br>(4) | 2007-2008<br>(5) |  |
| **Count, n (%)** | 8080 | 1516 (18.76) | 1550 (19.18) | 1669 (20.66) | 1419 (17.56) | 1926 (23.84) | <0.001 |
| **Age, mean (SD), yr** | 71.4 (7.8) | 71.1 (7.8) | 72.0 (8.0) | 72.3 (8.1) | 71.7 (8.1) | 70.4 (7.0) | <0.001 |
| **Age, n (%)** |  |  |  |  |  |  | <0.001 |
| 60-69 | 3619 | 716 (47.22) | 666 (42.97) | 694 (41.58) | 632 (44.54) | 911 (47.30) |  |
| 70-79 | 2669 | 496 (32.72) | 510 (32.90) | 551 (33.01) | 455 (32.06) | 657 (34.11) |  |
| 80-89 | 1792 | 304 (20.05) | 374 (24.13) | 424 (25.40) | 332 (23.40) | 358 (18.59) |  |
| **BLL, geometric mean (SE), µg/dl** | 2.18 (0.01) | 2.51 (0.04) | 2.21 (0.03) | 2.18 (0.03) | 2.19 (0.03) | 1.92 (0.02) | <0.001 |
| **Follow-up duration, mean (SD), yr** | 11.2 (5.3) | 12.9 (6.5) | 12.1 (5.9) | 11.1 (5.3) | 10.4 (4.5) | 9.6 (3.6) | <0.001 |
| **Status at end of follow-up, n (%)** |  |  |  |  |  |  | <0.001 |
| Death, Alzheimer's disease | 231 (2.86) | 45 (2.97) | 68 (4.39) | 54 (3.24) | 34 (2.40) | 30 (1.56) |  |
| Death, other | 4397 (54.42) | 1026 (67.68) | 930 (60.00) | 975 (58.42) | 696 (49.05) | 770 (39.98) |  |
| Alive | 3452 (42.72) | 445 (29.35) | 552 (35.61) | 640 (38.35) | 689 (48.56) | 1126 (58.46) |  |
| **Sex, n (%)** |  |  |  |  |  |  |  |
| Male | 4012 (49.65) | 756 (49.87) | 760 (49.03) | 817 (48.95) | 734 (51.73) | 945 (49.07) | 0.51 |
| Female | 4068 (50.35) | 760 (50.13) | 790 (50.97) | 852 (51.05) | 685 (48.27) | 981 (50.93) |  |
| **Race and ethnicity, n (%)** |  |  |  |  |  |  | <0.001 |
| Non-Hispanic White | 4676 (57.87) | 770 (50.79) | 957 (61.74) | 1002 (60.04) | 862 (60.75) | 1085 (56.33) |  |
| Non-Hispanic Black | 1387 (17.17) | 245 (16.16) | 252 (16.26) | 236 (14.14) | 292 (20.58) | 362 (18.80) |  |
| Hispanic | 1808 (22.38) | 471 (31.07) | 308 (19.87) | 375 (22.47) | 231 (16.28) | 423 (21.96) |  |
| Other | 209 (2.59) | 30 (1.98) | 33 (2.13) | 56 (3.36) | 34 (2.40) | 56 (2.91) |  |
| **Poverty-Index-Ratio, n (%)** |  |  |  |  |  |  | <0.001 |
| ≥2 | 3771 (46.67) | 585 (38.59) | 788 (50.84) | 790 (47.33) | 721 (50.81) | 887 (46.05) |  |
| <2 | 3538 (43.79) | 672 (44.33) | 634 (40.90) | 775 (46.43) | 608 (42.85) | 849 (44.08) |  |
| Missing | 771 (9.54) | 259 (17.08) | 128 (8.26) | 104 (6.23) | 90 (6.34) | 190 (9.87) |  |
| **Education, n (%)** |  |  |  |  |  |  | <0.001 |
| ≥12 years | 4812 (59.55) | 746 (49.21) | 950 (61.29) | 995 (59.62) | 900 (63.42) | 1221 (63.40) |  |
| <12 years | 3268 (40.45) | 770 (50.79) | 600 (38.71) | 674 (40.38) | 519 (36.58) | 705 (36.60) |  |
| **Current Smoker, n (%)** | 1344 (16.63) | 248 (16.36) | 244 (15.74) | 283 (16.96) | 262 (18.46) | 307 (15.94) | 0.27 |
| **History of CVD, n (%)** | 1919 (23.75) | 310 (20.45) | 351 (22.65) | 444 (26.60) | 355 (25.02) | 459 (23.83) | <0.001 |
| **History of cancer, n (%)** | 1478 (18.29) | 226 (14.91) | 307 (19.81) | 309 (18.51) | 264 (18.60) | 372 (19.31) | <0.01 |
| **History of diabetes, n (%)** | 1592 (19.70) | 282 (18.60) | 273 (17.61) | 331 (19.83) | 281 (19.80) | 425 (22.07) | <0.05 |
| **History of hypertension, n (%)** | 4490 (55.57) | 770 (50.79) | 810 (52.26) | 968 (58.00) | 808 (56.94) | 1134 (58.88) | <0.001 |
| **History of osteoporosis, n (%)** | 1021 (12.64) | 111 (7.32) | 200 (12.90) | 267 (16.00) | 170 (11.98) | 273 (14.17) | <0.001 |

Abbreviations: BLL, blood lead level.
\* Differences across cycles were tested using Chi-square or Kruskal-Wallis.

Consistent with secular declines in population-level lead exposure, geometric mean BLL declined steadily from 2.51 µg/dL in Cycle 1 to 1.92 µg/dL in Cycle 5, while mean follow-up decreased from 12.9 to 9.6 years. AD deaths peaked in Cycle 2 (n=68, 4.39%) and declined in later cycles (Cycle 5: n=30, 1.56%), partly reflecting shorter follow-up in recent cycles. The maximum BLL among AD decedents was only 8.4 µg/dL, substantially lower than the cohort maximum of 54 µg/dL.

In contrast to Horton et al., we found no association between log-transformed BLL and AD mortality in the pooled base model using updated mortality data (HR: 1.00; 95% CI: 0.78-1.28), likely reflecting the additional 150 AD deaths from extended follow-up. Cycle-stratified analyses revealed a temporal pattern (Supplemental Table S1), with associations shifting from inverse in early cycles (Cycle 1: HR=0.88, 95% CI: 0.51–1.52) to positive in later cycles (Cycle 5: HR=1.41, 95% CI: 0.71–2.80), though none reached statistical significance. In pooled-cycle models (Supplemental Table S2), calendar adjustment revealed associations obscured in crude analyses (unadjusted: HR=1.00, 95% CI: 0.78–1.28; calendar-adjusted: HR=0.81, 95% CI: 0.64–1.04). Excluding participants dying within 5 or 10 years strengthened this further (5-year: HR=0.77, 95% CI: 0.57–1.04; 10-year: HR=0.70, 95% CI: 0.49–1.01).

### Phase 2: NHANES 1988-2008

Of 27,085 eligible participants, we excluded 5,342 with BLL <1.0 µg/dL, 348 with missing follow-up data, and 87 exceeding top-coded age thresholds, yielding an analytic sample of 21,308 with 350 AD deaths over a median follow-up of 15.3 years (Figure 2). Participant characteristics by BLL category are presented in Table 2.

**Figure 2.**
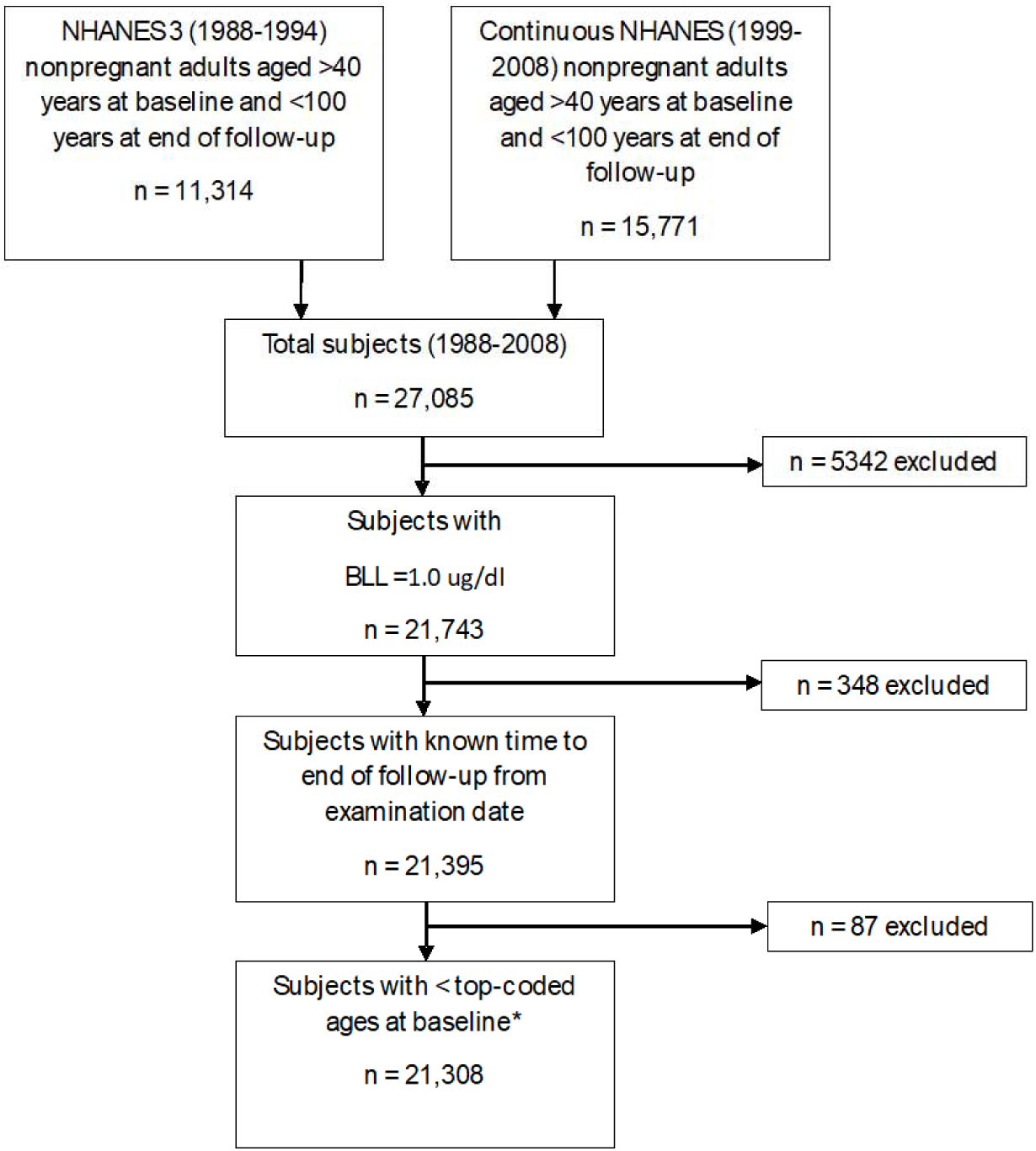
Flow chart.

**Table 2.**
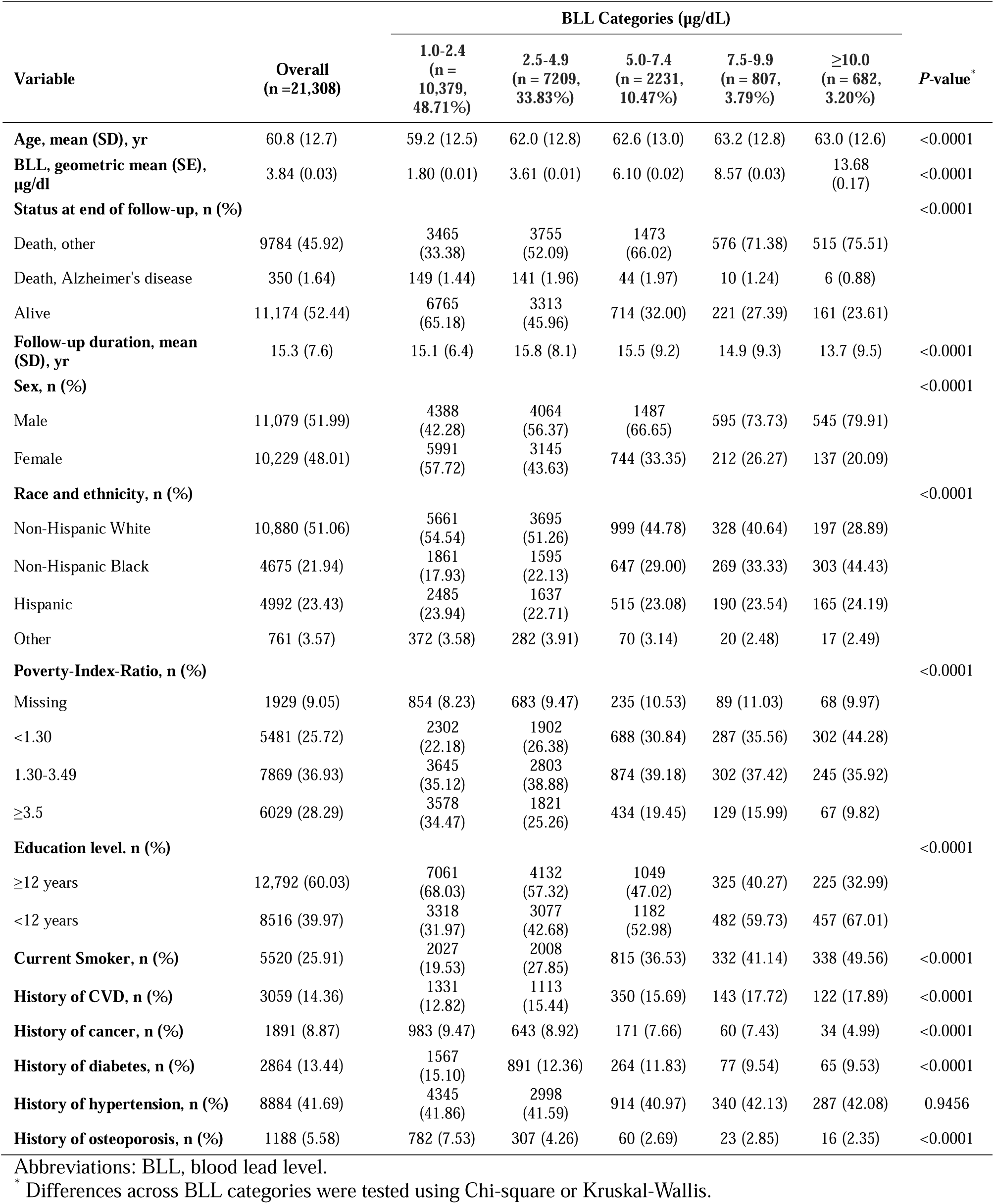
Demographic and clinical characteristics of combined NHANES III (1988-1994) and Continuous NHANES (1999-2008) study participants by BLL category.

| Variable | Overall<br>(n =21,308) | BLL Categories (µg/dL) |  |  |  |  | P-value* |
| --- | --- | --- | --- | --- | --- | --- | --- |
|  |  | 1.0-2.4<br>(n =<br>10,379,<br>48.71%) | 2.5-4.9<br>(n = 7209,<br>33.83%) | 5.0-7.4<br>(n = 2231,<br>10.47%) | 7.5-9.9<br>(n = 807,<br>3.79%) | ≥10.0<br>(n = 682,<br>3.20%) |  |
| Age, mean (SD), yr | 60.8 (12.7) | 59.2 (12.5) | 62.0 (12.8) | 62.6 (13.0) | 63.2 (12.8) | 63.0 (12.6) | <0.0001 |
| BLL, geometric mean (SE), µg/dl | 3.84 (0.03) | 1.80 (0.01) | 3.61 (0.01) | 6.10 (0.02) | 8.57 (0.03) | 13.68 (0.17) | <0.0001 |
| Status at end of follow-up, n (%) |  |  |  |  |  |  | <0.0001 |
| Death, other | 9784 (45.92) | 3465 (33.38) | 3755 (52.09) | 1473 (66.02) | 576 (71.38) | 515 (75.51) |  |
| Death, Alzheimer's disease | 350 (1.64) | 149 (1.44) | 141 (1.96) | 44 (1.97) | 10 (1.24) | 6 (0.88) |  |
| Alive | 11,174 (52.44) | 6765 (65.18) | 3313 (45.96) | 714 (32.00) | 221 (27.39) | 161 (23.61) |  |
| Follow-up duration, mean (SD), yr | 15.3 (7.6) | 15.1 (6.4) | 15.8 (8.1) | 15.5 (9.2) | 14.9 (9.3) | 13.7 (9.5) | <0.0001 |
| Sex, n (%) |  |  |  |  |  |  | <0.0001 |
| Male | 11,079 (51.99) | 4388 (42.28) | 4064 (56.37) | 1487 (66.65) | 595 (73.73) | 545 (79.91) |  |
| Female | 10,229 (48.01) | 5991 (57.72) | 3145 (43.63) | 744 (33.35) | 212 (26.27) | 137 (20.09) |  |
| Race and ethnicity, n (%) |  |  |  |  |  |  | <0.0001 |
| Non-Hispanic White | 10,880 (51.06) | 5661 (54.54) | 3695 (51.26) | 999 (44.78) | 328 (40.64) | 197 (28.89) |  |
| Non-Hispanic Black | 4675 (21.94) | 1861 (17.93) | 1595 (22.13) | 647 (29.00) | 269 (33.33) | 303 (44.43) |  |
| Hispanic | 4992 (23.43) | 2485 (23.94) | 1637 (22.71) | 515 (23.08) | 190 (23.54) | 165 (24.19) |  |
| Other | 761 (3.57) | 372 (3.58) | 282 (3.91) | 70 (3.14) | 20 (2.48) | 17 (2.49) |  |
| Poverty-Index-Ratio, n (%) |  |  |  |  |  |  | <0.0001 |
| Missing | 1929 (9.05) | 854 (8.23) | 683 (9.47) | 235 (10.53) | 89 (11.03) | 68 (9.97) |  |
| <1.30 | 5481 (25.72) | 2302 (22.18) | 1902 (26.38) | 688 (30.84) | 287 (35.56) | 302 (44.28) |  |
| 1.30-3.49 | 7869 (36.93) | 3645 (35.12) | 2803 (38.88) | 874 (39.18) | 302 (37.42) | 245 (35.92) |  |
| ≥3.5 | 6029 (28.29) | 3578 (34.47) | 1821 (25.26) | 434 (19.45) | 129 (15.99) | 67 (9.82) |  |
| Education level, n (%) |  |  |  |  |  |  | <0.0001 |
| ≥12 years | 12,792 (60.03) | 7061 (68.03) | 4132 (57.32) | 1049 (47.02) | 325 (40.27) | 225 (32.99) |  |
| <12 years | 8516 (39.97) | 3318 (31.97) | 3077 (42.68) | 1182 (52.98) | 482 (59.73) | 457 (67.01) |  |
| Current Smoker, n (%) | 5520 (25.91) | 2027 (19.53) | 2008 (27.85) | 815 (36.53) | 332 (41.14) | 338 (49.56) | <0.0001 |
| History of CVD, n (%) | 3059 (14.36) | 1331 (12.82) | 1113 (15.44) | 350 (15.69) | 143 (17.72) | 122 (17.89) | <0.0001 |
| History of cancer, n (%) | 1891 (8.87) | 983 (9.47) | 643 (8.92) | 171 (7.66) | 60 (7.43) | 34 (4.99) | <0.0001 |
| History of diabetes, n (%) | 2864 (13.44) | 1567 (15.10) | 891 (12.36) | 264 (11.83) | 77 (9.54) | 65 (9.53) | <0.0001 |
| History of hypertension, n (%) | 8884 (41.69) | 4345 (41.86) | 2998 (41.59) | 914 (40.97) | 340 (42.13) | 287 (42.08) | 0.9456 |
| History of osteoporosis, n (%) | 1188 (5.58) | 782 (7.53) | 307 (4.26) | 60 (2.69) | 23 (2.85) | 16 (2.35) | <0.0001 |

Abbreviations: BLL, blood lead level.
\* Differences across BLL categories were tested using Chi-square or Kruskal-Wallis.

Stratified analyses by 5-year exit intervals demonstrated substantial heterogeneity in the BLL-AD mortality association across calendar time, with hazard ratios ranging from 1.25 (95% CI: 0.45–3.47) in 1993–1997 to 0.54 (95% CI: 0.10–3.01) in 2018–2019 (Table 3), justifying calendar adjustment in pooled analyses. Maximum BLL among AD decedents was 13.3 µg/dL, lower than the cohort maximum of 56 µg/dL.

**Table 3.**
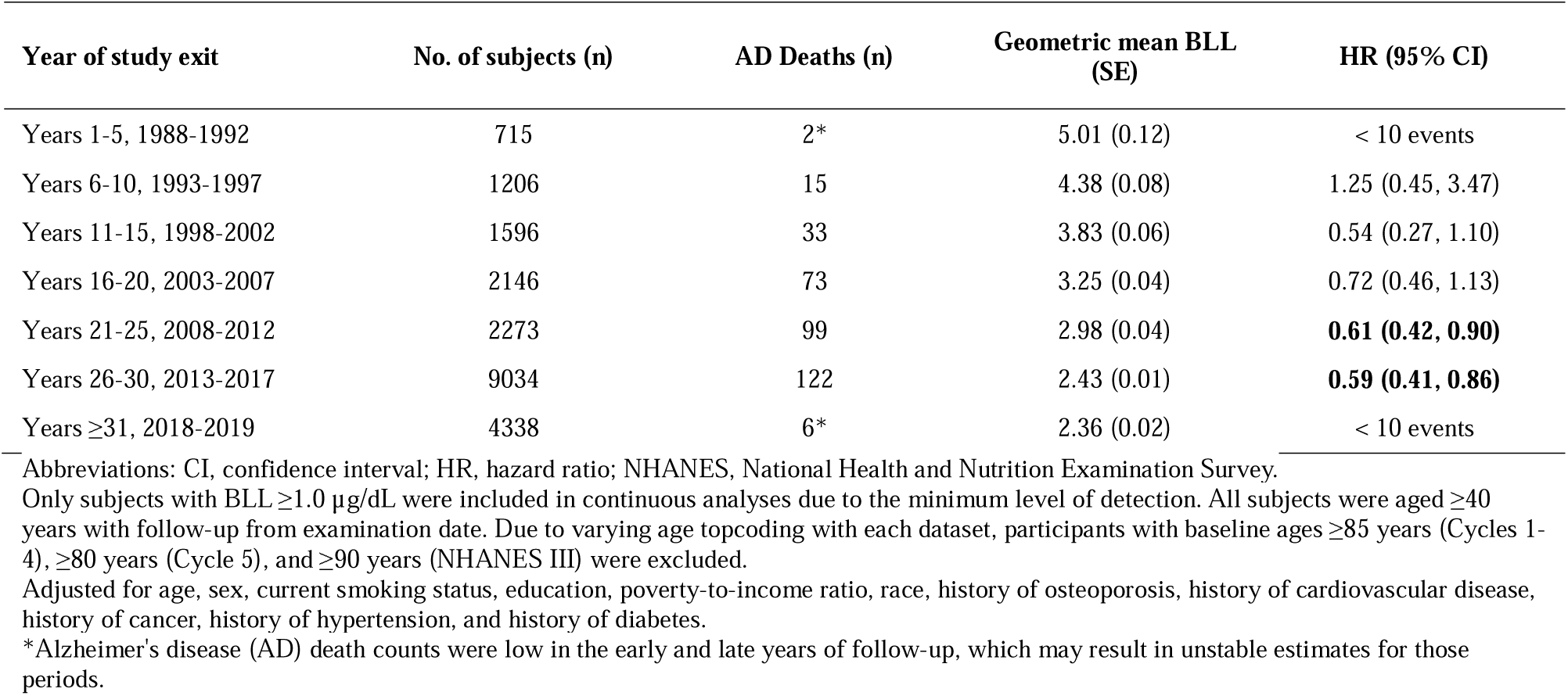
Natural log-transformed BLL as continuous predictor of AD mortality in NHANES III (1988-1994) and NHANES 1999-2008 combined data, stratified by year of study exit.

| Year of study exit | No. of subjects (n) | AD Deaths (n) | Geometric mean BLL (SE) | HR (95% CI) |
| --- | --- | --- | --- | --- |
| Years 1-5, 1988-1992 | 715 | 2* | 5.01 (0.12) | < 10 events |
| Years 6-10, 1993-1997 | 1206 | 15 | 4.38 (0.08) | 1.25 (0.45, 3.47) |
| Years 11-15, 1998-2002 | 1596 | 33 | 3.83 (0.06) | 0.54 (0.27, 1.10) |
| Years 16-20, 2003-2007 | 2146 | 73 | 3.25 (0.04) | 0.72 (0.46, 1.13) |
| Years 21-25, 2008-2012 | 2273 | 99 | 2.98 (0.04) | **0.61 (0.42, 0.90)** |
| Years 26-30, 2013-2017 | 9034 | 122 | 2.43 (0.01) | **0.59 (0.41, 0.86)** |
| Years ≥31, 2018-2019 | 4338 | 6* | 2.36 (0.02) | < 10 events |

Abbreviations: CI, confidence interval; HR, hazard ratio; NHANES, National Health and Nutrition Examination Survey.
Only subjects with BLL ≥1.0 µg/dL were included in continuous analyses due to the minimum level of detection. All subjects were aged ≥40 years with follow-up from examination date. Due to varying age topcoding with each dataset, participants with baseline ages ≥85 years (Cycles 1-4), ≥80 years (Cycle 5), and ≥90 years (NHANES III) were excluded.
Adjusted for age, sex, current smoking status, education, poverty-to-income ratio, race, history of osteoporosis, history of cardiovascular disease, history of cancer, history of hypertension, and history of diabetes.
\*Alzheimer's disease (AD) death counts were low in the early and late years of follow-up, which may result in unstable estimates for those periods.

In pooled analyses (Table 4), unadjusted models showed a nonsignificant inverse association (HR=0.86, 95% CI: 0.70–1.05, p=0.13). After calendar adjustment, the association strengthened substantially (HR=0.57, 95% CI: 0.46–0.70, p<0.001) and persisted after excluding early deaths (5-year: HR=0.55, 95% CI: 0.44–0.68; 10-year: HR=0.54, 95% CI: 0.42–0.69; both p<0.001).

**Table 4.**
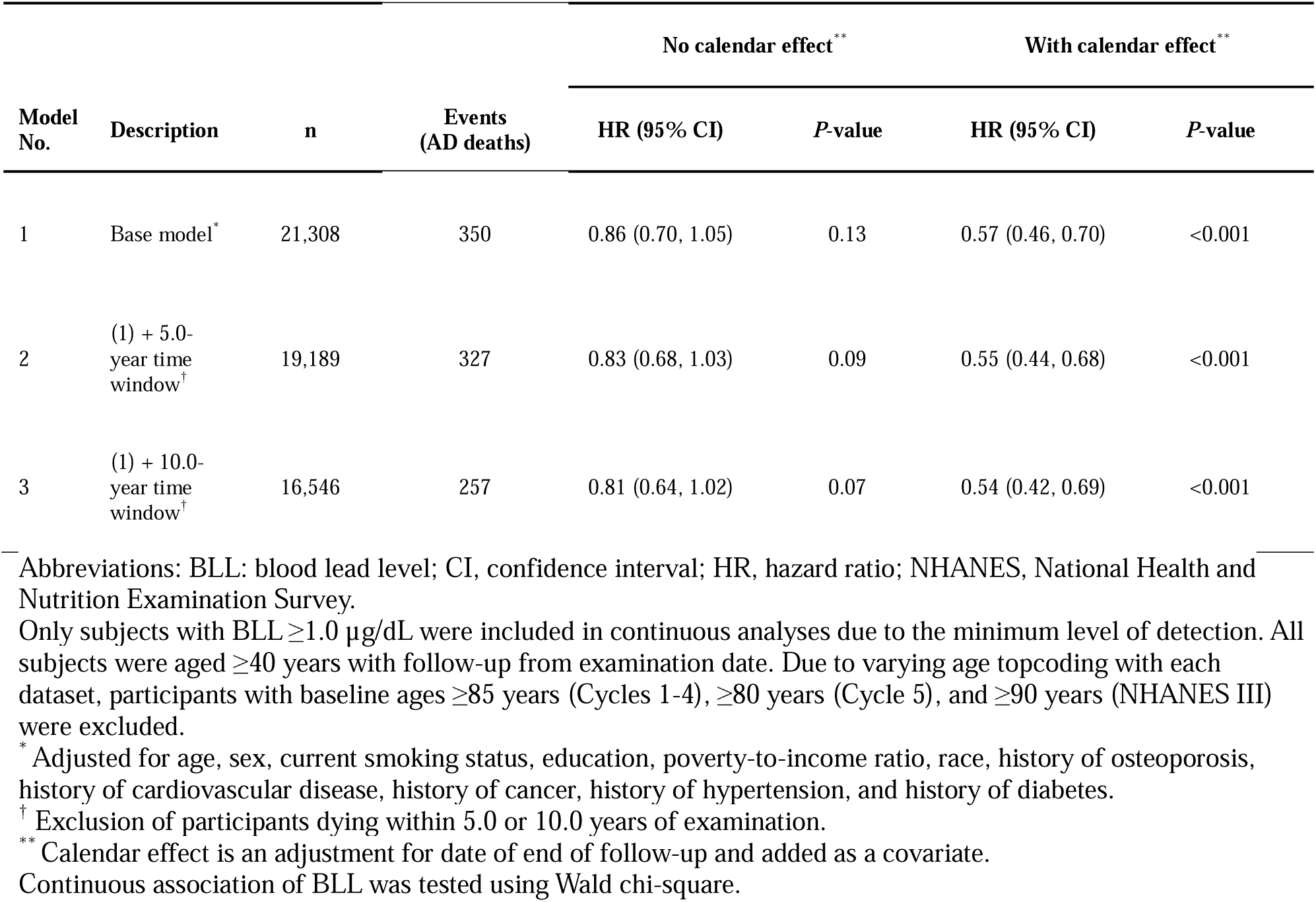
Natural log-transformed BLL as continuous predictor of AD mortality in NHANES III (1988-1994) and NHANES 1999-2008 combined data, adjusted with and without calendar effect covariate.

| Model No. | Description | n | Events (AD deaths) | No calendar effect** |  | With calendar effect** |  |
| --- | --- | --- | --- | --- | --- | --- | --- |
|  |  |  |  | HR (95 % CI) | P-value | HR (95% CI) | P-value |
| 1 | Base model* | 21,308 | 350 | 0.86 (0.70, 1.05) | 0.13 | 0.57 (0.46, 0.70) | <0.001 |
| 2 | (1) + 5.0-year time window<sup>†</sup> | 19,189 | 327 | 0.83 (0.68, 1.03) | 0.09 | 0.55 (0.44, 0.68) | <0.001 |
| 3 | (1) + 10.0-year time window<sup>†</sup> | 16,546 | 257 | 0.81 (0.64, 1.02) | 0.07 | 0.54 (0.42, 0.69) | <0.001 |

Abbreviations: BLL: blood lead level; CI, confidence interval; HR, hazard ratio; NHANES, National Health and Nutrition Examination Survey.
Only subjects with BLL $\geq 1.0$ $\mu\text{g/dL}$ were included in continuous analyses due to the minimum level of detection. All subjects were aged $\geq 40$ years with follow-up from examination date. Due to varying age topcoding with each dataset, participants with baseline ages $\geq 85$ years (Cycles 1-4), $\geq 80$ years (Cycle 5), and $\geq 90$ years (NHANES III) were excluded.
\* Adjusted for age, sex, current smoking status, education, poverty-to-income ratio, race, history of osteoporosis, history of cardiovascular disease, history of cancer, history of hypertension, and history of diabetes.
<sup>†</sup> Exclusion of participants dying within 5.0 or 10.0 years of examination.
\*\* Calendar effect is an adjustment for date of end of follow-up and added as a covariate.
Continuous association of BLL was tested using Wald chi-square.

Categorical analyses revealed a strong monotonic inverse dose-response after calendar adjustment (Table 5): compared to the reference category (1.0–2.4 µg/dL), hazard ratios decreased progressively across higher BLL categories (2.5–4.9: HR=0.69; 5.0–7.4: HR=0.56; 7.5–9.9: HR=0.36; ≥10: HR=0.31; p-trend<0.001), persisting after excluding early deaths. Spline analyses corroborated these findings, showing a strong monotonic inverse relationship after calendar adjustment with the steepest gradient between 1.5–7 µg/dL, plateauing above 10 µg/dL (Figure 3).

**Figure 3.**
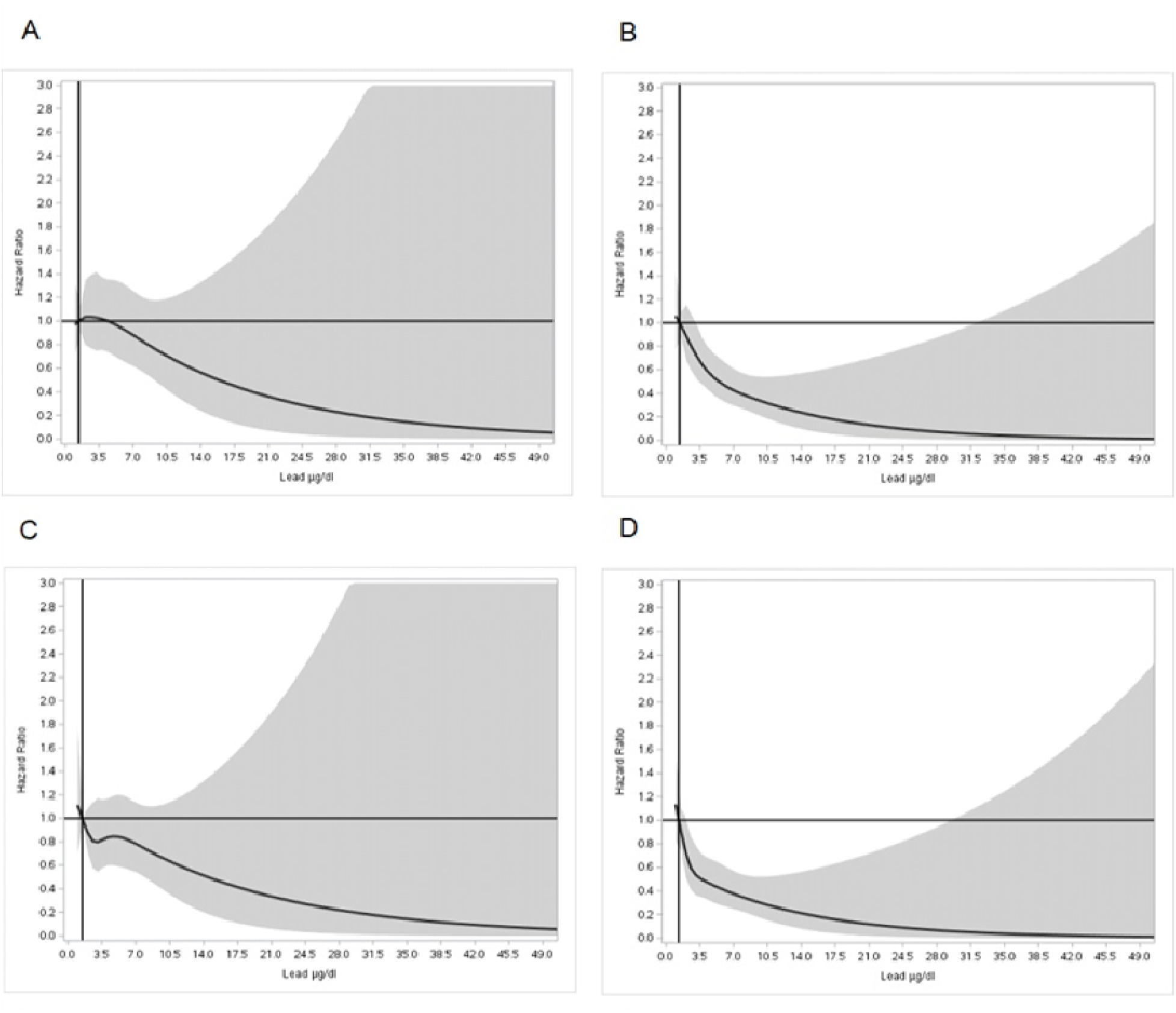
Restricted cubic splines of the association of blood lead level and subsequent Alzheimer’s disease (AD) mortality. Hazard ratios (solid lines) and 95% confidence intervals (shaded regions) from Cox proportional hazards models with restricted cubic splines (5 knots at 5th, 27.5th, 50th, 72.5th, and 95th percentiles). The reference value (HR=1.0) is set at the 12.5th percentile of the blood lead distribution (1.4 µg/dL). All models adjust for age, sex, race/ethnicity, education, poverty-income ratio, smoking status, and medical history (cardiovascular disease, cancer, hypertension, diabetes, osteoporosis). **Panel A:** Full sample (n=21,308) without calendar adjustment. **Panel B:** Full sample with calendar adjustment for study exit time. **Panel C:** Participants with ≥10 years of follow-up (excluding deaths within first 10 years) without calendar adjustment. **Panel D:** Participants with ≥10 years of follow-up (excluding deaths within first 10 years) with calendar adjustment.

**Table 5.**
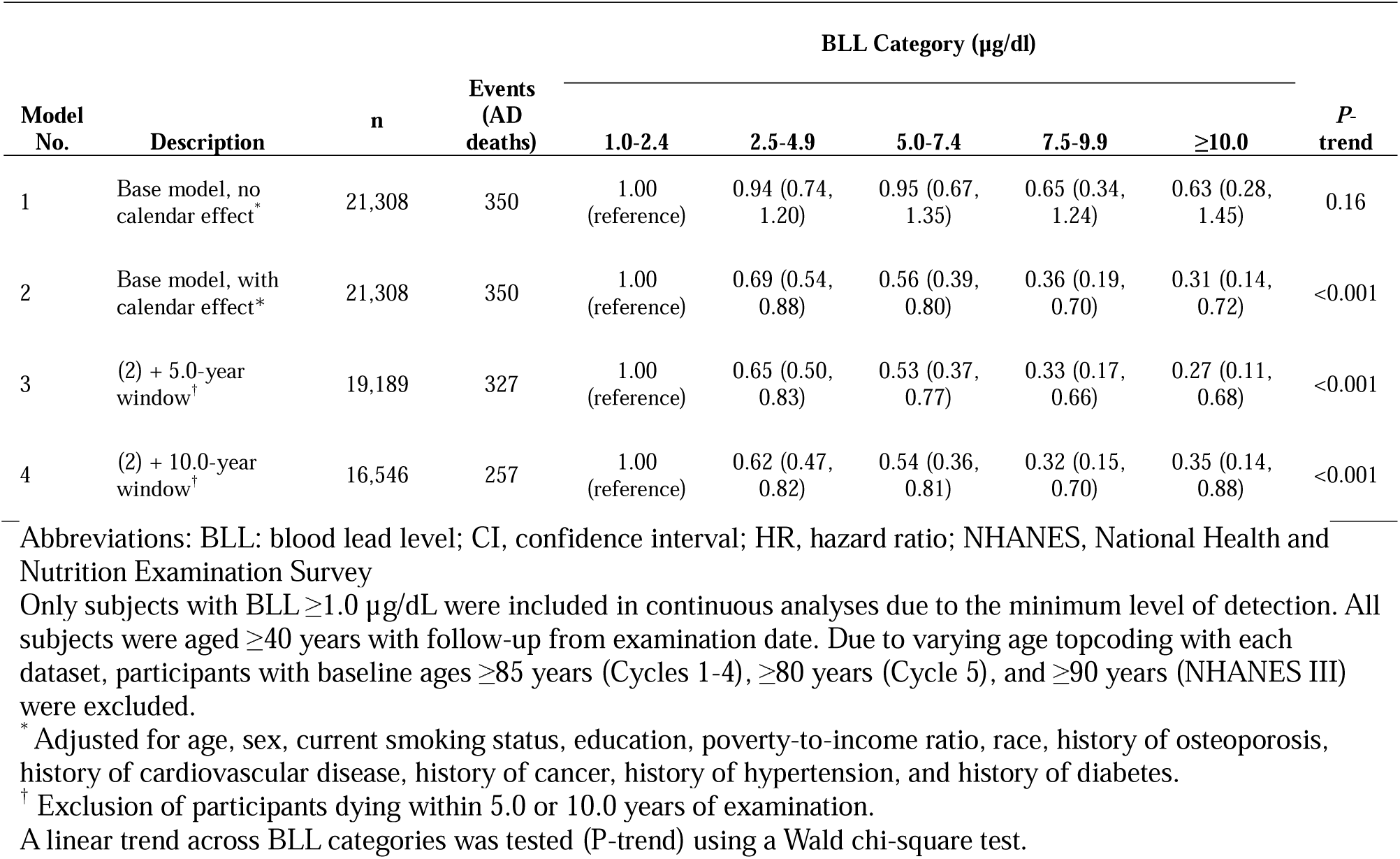
Adjusted HRs (95% CI) of categorical BLL predicting AD mortality in NHANES III (1988-1994) and NHANES 1999-2008 combined data.

Calendar adjustment notably altered demographic associations. Before adjustment, non-Hispanic Black and Hispanic participants appeared to have lower AD mortality than non-Hispanic Whites, consistent with prior national mortality reports. After calendar adjustment, these associations disappeared entirely, suggesting they reflected differential follow-up and exposure histories rather than true biological differences. Cancer history was strongly associated with increased AD mortality, strengthening after calendar adjustment. Full covariate results are presented in Supplementary Table S3.

To evaluate competing mortality, we compared age at death across BLL categories (Supplementary Table S4). Among other-cause decedents, mean age at death was remarkably consistent across the three lowest BLL categories (79.0, 79.8, and 79.1 years), which accounted for 95.4% (334/350) of all AD deaths. This suggests that the concentration of AD mortality in lower BLL groups is not driven by earlier death from competing causes. Mean age at other-cause death was only modestly lower in the highest BLL categories (78.4 and 76.9 years), which contributed only 16 AD deaths. While age at study exit among survivors was lower (71.6–77.1 years), this reflects the recruitment of survivors from later survey cycles with shorter follow-up—the same temporal confounding addressed by our primary analysis.

## Discussion

We hypothesized that declining population-level lead exposure over recent decades may contribute to rising AD incidence through competitive inhibition of other neurotoxic metals. This hypothesis was motivated by three observations: (1) age-standardized AD mortality in the U.S. nearly doubled as BLL declined by more than 50% between 1999 and 2016,[24, 25] (2) international studies show lower AD prevalence in countries with higher environmental lead exposure,[26–28] and (3) metals implicated in AD pathogenesis share common transport pathways through which competitive inhibition could occur, with this mechanism being supported by complementary observations in neurodevelopment.[4, 5, 29, 30]

Using extended NHANES mortality follow-up data spanning 1988-2019 for the Phase 1 Continuous NHANES dataset, we found that temporal confounding strongly influenced the apparent BLL–AD mortality association. In conventionally adjusted pooled analyses, the association appeared null, markedly differing from the positive, though nonsignificant, association reported by Horton et al. (HR=1.28, 95% CI: 0.97-1.70). This attenuation likely reflects the 150 additional AD deaths from extended follow-up, which redistributed mortality across cycles in ways that diluted the enrollment-driven ascertainment imbalance driving Horton’s positive estimate.[6] Cycle-stratified analyses revealed marked heterogeneity: hazard ratios varied from inverse in early survey cycles to positive in later cycles, despite declining population-level lead exposure. After adjusting for calendar time at study exit—which accounts for differential exposure trajectories and unequal ascertainment opportunity—a robust inverse dose-response relationship emerged.

In the Phase 2 combined sample (n=21,308), each unit increase in log-transformed BLL corresponded to a 43% lower AD mortality risk (HR = 0.57, 95% CI: 0.46–0.70). This transformation from null to strongly inverse after calendar adjustment highlights how traditional survival models can obscure or even reverse associations when both exposure and outcome ascertainment vary across calendar time. Moreover, the temporal stratification analyses exhibited features characteristic of Simpson’s paradox, with associations within calendar-specific strata differing substantially from the aggregate estimate without the added study exit covariate.

The temporal confounding we identified reflects three interconnected mechanisms. First, secular exposure declines mean that participants recruited earlier had both higher baseline BLLs and greater subsequent declines. Standard survival models that adjust only for baseline BLL cannot distinguish between these different cumulative exposure scenarios. Second, AD mortality is only observable among decedents, and earlier cohorts with higher BLL and longer observation periods have substantially greater opportunity for such a death compared to later cohorts, creating a spurious positive association. Third, period effects including changes in diagnostic practices and cause-of-death coding systematically vary across calendar years and cannot be removed by adjusting for survey cycle or baseline year alone.

The calendar effect covariate addresses these issues by effectively comparing participants who exited the study at the same calendar time, equalizing both exposure trajectories and ascertainment opportunities. The primary driver of the adjustment’s impact was not the treatment of deaths per se, but how survivors were handled: assigning all living participants a common exit date eliminated the systematic bias in which later, lower-exposure participants were disproportionately classified as non-events. The inverse relationship was robust across sensitivity analyses excluding early deaths to minimize potential reverse causality. [22, 23]

Competition among metals for biological transport may influence neurological outcomes.[29, 31] While AD differs clinically from neurodevelopmental disorders, similar competitive interactions could plausibly affect neurodegenerative processes. Under such circumstances, population-level shifts in exposure to one metal could alter the bioavailability of others, potentially producing counterintuitive exposure–disease relationships. Our hypothesized mechanism—that lead competitively inhibits the transport or bioavailability of other divalent metals implicated in AD pathogenesis—provides a biologically plausible explanation for the inverse association observed in the current study. The divalent metal transporter-1 (DMT-1) shows hierarchical metal selectivity, raising the possibility that higher lead levels could reduce brain accumulation of metals that may be involved in oxidative damage,[30, 32] particularly iron, copper, and zinc.[4, 5] Under this model, declining environmental lead would increase bioavailability of AD-relevant metals, thereby elevating disease risk. While competitive inhibition is a compelling framework, it remains speculative without targeted validation. Specifically, research must (1) demonstrate that lead inhibits transport of AD-relevant metals; (2) employ animal models to show that reduced brain levels of these metals attenuate AD pathology; and (3) utilize longitudinal human studies linking serial metal measurements to incident AD diagnosis.

Calendar adjustment altered demographic associations in revealing ways. The apparent protective effects of non-Hispanic Black and Hispanic ethnicity on AD mortality disappeared after adjustment, suggesting they reflected differential follow-up and exposure histories rather than true biological differences. Sex differences strengthened after adjustment, with women showing lower AD mortality than men, consistent with longer survival following dementia diagnosis.[33] Cancer history was associated with increased AD mortality; an association that strengthened after calendar adjustment. This is consistent with prior literature: though studies have found early decreased risk for cancer survivors, risk appears to return to null or even increase among longer-term survivors (≥5–10 years).[34–36]

Limited prior epidemiologic evidence exists on BLL and AD. An Iranian case-control study reported higher BLL among AD cases, but this finding was heavily confounded by opium use (BLL in opium-using AD patients: 54.5 µg/dL vs. 3.8 µg/dL in non-users).[8] Hare et al. found no serum lead differences between AD patients and controls, though their cross-sectional design precluded assessment of temporality, including reverse causality.[7] The aforementioned study by Wang et al. found no association of BLL with incident AD; however, temporal confounding was not addressed, and reverse causality arising from physiological redistribution of lead near the time of diagnosis was not examined.[9] Further, their heterogeneous AD ascertainment approach, incorporating both incidence from Medicare claims and mortality via the NDI, was not disaggregated by source, leaving the structural confounding illustrated here unknown. Critically, the strong positive association Wang et al. observed with bone lead may itself be a product of this confounding. Unlike blood lead, which responds quite rapidly to changes in exposure, bone lead reflects cumulative exposure, making bone lead even more intrinsically linked to the temporal structure of enrollment than BLL.[37] Earlier cohorts, having lived through peak environmental lead levels before regulatory decline, would have entered the study with substantially higher bone lead burdens than later cohorts and would contribute disproportionate follow-up time and ascertainment opportunity, amplifying rather than attenuating the structural confounding described here.[38]

Studies of lead and cognitive function show heterogeneous findings: associations are consistent at occupational exposure levels (often >40 µg/dL), but an EPA review identified no clear relationship in general population studies with lower exposures.[39, 40] It is critical to distinguish between cognitive impairment and AD. The former may be a precursor of the latter; however, this is not invariably so.[41] In individuals exposed to extreme levels of lead, particularly in the occupational setting, cognitive impairment may arise from an alternative mechanism.

At the population level, striking geographic disparities in AD prevalence, with markedly lower rates in Nigeria and rural India compared to the United States, are consistent with environmental contributions to AD risk.[26, 27] While diagnostic standardization across regions presents methodological challenges, these carefully designed studies employed culturally adapted instruments and validated diagnostic criteria. Global variation in lead exposure could potentially have contributed to these differences.[25, 28, 42] This hypothesis remains speculative, however, and is complicated by long latency between exposure and disease onset and by challenges in diagnostic standardization across settings.

This study has important limitations. First, the outcome reflects Alzheimer’s disease (AD) mortality rather than incidence, and AD is likely under-ascertained on death certificates, which capture only the primary cause of death.[43] In addition, diagnostic recognition and certification of AD may have improved over the study period. However, this potential bias operates through calendar time rather than through a direct association between BLL and diagnostic accuracy. It is therefore addressed by inclusion of the calendar exit covariate, which standardizes outcome ascertainment opportunity across time periods. For residual confounding to explain our findings, improved ascertainment would need to differentially favor lower-BLL individuals within the same calendar period, a mechanism for which there is no clear biological or administrative basis. Consistent with this interpretation, stratified analyses by exit year show that the inverse association is already present at an early period (1998–2002; HR = 0.54) and remains relatively stable across subsequent periods, rather than strengthening over time as would be expected if secular improvements in ascertainment were driving the association.

Second, single baseline BLL measures cannot fully capture lifetime exposure trajectories, though this limitation is standard in large epidemiologic cohorts.[44–48] Moreover, non-differential misclassification would bias hazard ratios toward the null (HR = 1.0), suggesting our observed inverse association (HR=0.57) may underestimate the true magnitude.[49] Third, the calendar covariate simplifies complex temporal processes into a single adjustment term and has not been validated in independent, long-latency datasets. However, the consistency of findings across exit-year strata and sensitivity analyses provides empirical support for its validity in this setting. Finally, while competing mortality is a classic concern in late-life cohort analyses, it does not explain the observed inverse associations. Within the BLL categories encompassing 95.4% (334/350) of all observed AD deaths (1.0 to 7.4 µg/dL, Table S4), the mean age at other-cause mortality was remarkably uniform, ranging from 79.0 to 79.8 years. This stability in age at competing death implies that participants in low-exposure groups were not simply at risk for AD longer than those in higher-lead groups.

These findings should not alter current public health policy on lead exposure. Lead remains a well-established neurotoxin with documented adverse effects, particularly on child development. Any observed inverse association with AD mortality—if not explained by confounding, competing mortality, or methodological artifacts—would require extensive replication and mechanistic validation before informing policy decisions.

Our study has important methodological implications for environmental epidemiology. We demonstrate that failure to account for secular trends in exposure and differential mortality-dependent outcome ascertainment may obscure or reverse exposure-disease associations in long-term studies. The calendar adjustment approach offers a generalizable framework: by modeling exit time explicitly, it corrects differential outcome ascertainment and equalizes opportunity for event detection across participants. Future work should validate this method in other cohorts with long-latency outcomes.

## Conclusion

After adjusting for calendar time at study exit—a method that equalizes exposure trajectories and outcome ascertainment opportunities—we observed a strong, inverse association between BLL and AD mortality that was not apparent in standard analyses. This study demonstrates that temporal confounding can profoundly distort exposure-disease associations in long-term epidemiologic investigations. Whether this inverse relationship reflects competitive metal interactions, competing risks, or residual confounding cannot be determined from observational data alone and requires experimental validation. Importantly, our results do not support any beneficial role for lead exposure and should not influence current public health policy. Lead’s well-established toxicity justifies continued efforts to minimize population exposure.

## Statements

### Ethics approval and consent to participate

Study protocols for NHANES were approved by the NCHS Ethics Review Board. All adult participants provided signed informed consent.

### Availability of data and materials

All data can be found on the CDC’s National Center for Health Statistics webpage: https://www.cdc.gov/nchs/nhanes/index.htm. SAS code is available from the author upon request.

### Authors’ contributions

The author was the sole contributor to all aspects of the study and manuscript development.

### Artificial Intelligence

The author used large language model AI assistants to support manuscript development and editing. All AI-generated content was reviewed, verified, and revised by the author, who takes full responsibility for the accuracy and integrity of the work.

