## Supplement Tables for "Blood Lead Levels and Alzheimer’s Disease Mortality in NHANES: Addressing Temporal Confounding Through a Study Exit Covariate"

**Supplementary Tables**

**Table S1.** Comparison of HRs (95% CI) of natural log-transformed BLL predicting AD mortality in Continuous NHANES (1999-2008), by NHANES cycles.

| **Model**  **No.** | **Description** | **Cycle 1,**  **1999-2000** | **Cycle 2,**  **2001-2002** | **Cycle 3,**  **2003-2004** | **Cycle 4,**  **2005-2006** | **Cycle 5,**  **2007-2008** |
| --- | --- | --- | --- | --- | --- | --- |
| 1 | Base model* | 0.88 (0.51, 1.52), n = 1516 | 0.94 (0.59, 1.49), n = 1550 | 1.17 (0.69, 1.98), n = 1669 | 0.90 (0.46, 1.75), n = 1419 | 1.41 (0.71, 2.80), n = 1926 |
| 2 | (1) + Modified criteria^‡^ | 0.82 (0.48, 1.41), n = 1935 | 1.09 (0.65, 1.84), n = 2036 | 1.24 (0.65, 2.37), n = 2040 | 0.91 (0.42, 1.97), n = 1880 | 1.41 (0.54, 3.67), n = 2417 |
| 3 | (2) + 5.0-year window^¶^ | 0.81 (0.46, 1.41), n = 1719 | 0.99 (0.57, 1.74), n = 1828 | 1.23 (0.62, 2.44), n = 1779 | 0.64 (0.27, 1.54), n = 1678 | 1.18 (0.43, 3.26), n = 2232 |
| 4 | (2) + 10.0-year window^¶^ | 0.85 (0.46, 1.58), n = 1418 | 1.00 (0.52, 1.94), n = 1534 | 0.78 (0.33, 1.81), n = 1495 | 0.37 (0.12, 1.19), n = 1426 | 1.01 (0.23, 4.43), n = 1947 |

Abbreviations: BLL: blood lead level; CI, confidence interval; HR, hazard ratio; NHANES, National Health and Nutrition Examination Survey; AD: Alzheimer’s disease

^*^Included all subjects aged ≥60 years. Adjusted for age, sex, poverty status, race/ethnicity, and smoking status.

^‡^ BLL ≥0.3 µg/dL, adults ≥50 years, < top-coded age (exclusion of participants aged 85 and 80 years from Cycles 1-4 and Cycle 5, respectively), additional covariates (education, history of osteoporosis, history of CVD, history of hypertension, history of cancer, and history of diabetes).

^¶^ Exclusion of participants dying within 5.0 or 10.0 years of examination.

**Table S2.** Comparison of HRs (95% CI) of natural log-transformed BLL predicting AD mortality in Continuous NHANES (1999-2008) pooled cycles, with and without calendar effect adjustment.

| **Model No.** | **Description** | **n** | **Events**  **(AD deaths)** | **No calendar effect^**^** | | **With calendar effect^**^** | |
| --- | --- | --- | --- | --- | --- | --- | --- |
|  |  |  |  | **HR (95% CI)** | ***P*-value** | **HR (95% CI)** | ***P*-value** |
| 1 | Base model^*^ | 8080 | 231 | 1.00 (0.78, 1.28) | 0.97 | 0.81 (0.64, 1.04) | 0.10 |
| 2 | (1) + Modified criteria^‡^ | 10,308 | 186 | 0.99 (0.76, 1.31) | 0.96 | 0.84 (0.63, 1.10) | 0.21 |
| 3 | (2) + 5.0-year window^¶^ | 9236 | 166 | 0.91 (0.68, 1.22) | 0.53 | 0.77 (0.57, 1.04) | 0.09 |
| 4 | (2) + 10.0-year window^¶^ | 7820 | 116 | 0.79 (0.56, 1.13) | 0.19 | 0.70 (0.49, 1.01) | 0.05 |

Abbreviations: BLL: blood lead level; CI, confidence interval; HR, hazard ratio; NHANES, National Health and Nutrition Examination Survey; AD: Alzheimer’s disease

^*^Included all subjects aged ≥60 years. Adjusted for age, sex, poverty status, race/ethnicity, and smoking status.

^‡^ BLL ≥0.3 µg/dL, adults ≥50 years, < top-coded age (exclusion of participants aged 85 and 80 years from Cycles 1-4 and Cycle 5, respectively), additional covariates (education, history of osteoporosis, history of CVD, history of hypertension, history of cancer, and history of diabetes).

^¶^ Exclusion of participants dying within 5.0 or 10.0 years of examination.

^**^Calendar effect covariate represents approximate calendar time at study exit, calculated by adding each participant's follow-up time (months from examination to exit) to a cycle-specific offset (0, 24, 48, 72, and 96 months for Cycles 1–5, respectively), and was included as a continuous predictor in Cox models.

Continuous association of BLL was tested using Wald chi-square.

**Table S3**. Mutually adjusted hazard ratios for covariates in relation to Alzheimer’s disease mortality: Analysis of natural log-transformed blood lead levels as a continuous predictor in combined NHANES III (1988-1994) and NHANES 1999-2008 Data

|  | **No calendar effect**^**^ | | **With calendar effect**^**^ | |
| --- | --- | --- | --- | --- |
| **Covariate** | **HR (95% CI)** | ***P*-value** | **HR (95% CI)** | ***P*-value** |
| **Age (per 1-year increment)** | 1.19 (1.17, 1.21) | <0.0001 | 1.16 (1.14, 1.17) | <0.0001 |
| **Sex (male)** |  |  |  |  |
| Female | 0.94 (0.75, 1.18) | 0.59 | 0.82 (0.65, 1.03) | 0.09 |
| **Race/Ethnicity (Non-Hispanic White)** |  |  |  |  |
| Non-Hispanic Black | 0.86 (0.62, 1.19) | 0.36 | 1.00 (0.72, 1.40) | 0.99 |
| Hispanic | 0.80 (0.59, 1.09) | 0.15 | 1.03 (0.76, 1.41) | 0.85 |
| Other | 0.78 (0.41, 1.48) | 0.45 | 1.01 (0.53, 1.92) | 0.98 |
| **Education (Low)** |  |  |  |  |
| High | 0.93 (0.73, 1.18) | 0.54 | 1.09 (0.86, 1.39) | 0.48 |
| **Poverty-Income-Ratio (0-1.3)** |  |  |  |  |
| Missing | 0.84 (0.57, 1.24) | 0.39 | 0.82 (0.55, 1.21) | 0.31 |
| 1.3-3.48 | 0.91 (0.69, 1.21) | 0.51 | 0.89 (0.67, 1.17) | 0.39 |
| 3.49 | 0.78 (0.55, 1.11) | 0.17 | 0.77 (0.54, 1.09) | 0.13 |
| **Smoking Status (No)** |  |  |  |  |
| Yes | 0.99 (0.72, 1.37) | 0.97 | 0.91 (0.66, 1.25) | 0.56 |
| **Osteoporosis (No)** |  |  |  |  |
| Yes | 0.93 (0.62, 1.39) | 0.71 | 1.17 (0.78, 1.75) | 0.45 |
| **Cardiovascular disease (No)** |  |  |  |  |
| Yes | 1.16 (0.86, 1.57) | 0.33 | 1.10 (0.81, 1.48) | 0.54 |
| **Cancer (No)** |  |  |  |  |
| Yes | 1.58 (1.16, 2.14) | <0.01 | 1.86 (1.37, 2.52) | <0.0001 |
| **Hypertension (No)** |  |  |  |  |
| Yes | 0.89 (0.71, 1.11) | 0.29 | 0.97 (0.78, 1.21) | 0.81 |
| **Diabetes (No)** |  |  |  |  |
| Yes | 1.06 (0.75, 1.48) | 0.75 | 0.96 (0.68, 1.34) | 0.80 |
| **Calendar Effect**^*^ **(per 1-month increment)** | - | - | 0.99 (0.99, 0.99) | <0.0001 |

Only subjects with BLL ≥1.0 µg/dL were included in continuous analyses due to the minimum level of detection. All subjects were aged ≥40 years with follow-up from examination date. Maximum age of subjects: NHANES III, <90; NHANES Cycles 1-4, <85; and NHANES Cycle 5 <80.

^*^ Calendar effect is an adjustment for date of end of follow-up and added as a covariate.

**Table S4.** Age at study exit by blood lead level category and outcome group, NHANES 1988–2008

|  | **AD Deaths** | | | **Other-Cause Deaths** | | | **Alive at End of Follow-up** | | |
| --- | --- | --- | --- | --- | --- | --- | --- | --- | --- |
| **BLL Category (µg/dL)** | **N** | **Mean (SD)** | **Median** | **N** | **Mean (SD)** | **Median** | **N** | **Mean (SD)** | **Median** |
| 1.0–2.4 | 149 | 86.1 (6.8) | 87.4 | 3,465 | 79.0 (11.2) | 80.9 | 6,765 | 71.6 (10.2) | 70.8 |
| 2.5–4.9 | 141 | 87.4 (6.8) | 88.8 | 3,755 | 79.8 (10.7) | 81.7 | 3,313 | 75.1 (9.8) | 74.3 |
| 5.0–7.4 | 44 | 86.7 (6.5) | 87.5 | 1,473 | 79.1 (10.9) | 80.6 | 714 | 75.8 (9.3) | 75.3 |
| 7.5–9.9 | 10 | 81.3 (8.1) | 81.8 | 576 | 78.4 (10.7) | 79.4 | 221 | 77.1 (9.0) | 76.3 |
| ≥10.0 | 6 | 86.8 (4.2) | 87.9 | 515 | 76.9 (10.8) | 78.3 | 161 | 76.0 (8.8) | 76.0 |

Age at study exit is age at death for decedents and age at end of follow-up (December 2019) for survivors. BLL, blood lead level. Values are mean (standard deviation) and median age in years.
